# Global Measurement of Physical Intimate Partner Violence to Monitor Sustainable Development Goal 5

**DOI:** 10.1101/2021.07.01.21259594

**Authors:** Kathryn M. Yount, Yuk Fai Cheong, Zara Khan, Irina Bergenfeld, Nadine Kaslow, Cari Jo Clark

**Affiliations:** Hubert Department of Global Health, Rollins School of Public Health, Emory University, 1518 Clifton Rd, NE, Room 7029, Atlanta, GA 30322; Department of Sociology, Emory University, 1555 Dickey Drive; Department of Psychology, Emory University, 36 Eagle Row, Atlanta, GA 30322; University of Texas Southwestern Medical School, 5323 Harry Hines Blvd, Dallas, TX 75390; Department of Psychiatry and Behavioral Sciences, Emory University School of Medicine, 12 Executive Park Dr, Atlanta, GA 30329

## Abstract

**Background:** One third of women experience IPV and potential sequelae. Sustainable Development Goal (SDG) 5.2—to eliminate all violence against women, including IPV— compels national governments to monitor such violence. We conducted the first global measurement-invariance assessment of standardized physical IPV items.

**Methods:** Thirty-six Demographic and Health Surveys (DHS) from 36 Lower-/Middle-Income Countries (LMICs) administering the same 18 IPV items during 2012-2018 were included. We performed exploratory and confirmatory factor analyses (EFA/CFA) with seven physical IPV items, which are the most behaviorally specific and reliable. Datasets meeting EFA/CFA model fit criteria (loadings>.35, RMSEA<.08, CFI/TLI>.95) were included in multiple-group CFA to test strict measurement invariance, and in alignment optimization (AO) to test approximate measurement invariance. We compared national rankings based on AO-derived scores and lifetime physical IPV prevalences, and correlated AO-dervied scores with physical, sexual, and psychological IPV prevalences.

**Results:** Estimated lifetime physical IPV varied widely (5.6%-50.5%). All loadings and fit statistics met thresholds in country-specific EFA/CFAs. A unidimensional, seven-item physical IPV construct lacked scalar invariance in multiple-group CFA but achieved approximate measurement invariance in AO analysis, as 12.3% (<25%) of model parameters were non-invariant. National rankings of AO-derived scores and estimated physical IPV prevalences were similarly distributed, but national estimates often were not significantly different, so grouped score ranges or prevalence ranges are advised. Three items (slap, twist, choke) warrant cognitive testing to improve their psychometric performance. Correlations of AO-derived scores with IPV prevalences ranged from .48 to .66.

**Conclusions:** Seven DHS physical-IPV items were approximately invariant across 36 LMICs spanning five regions and are reasonable for cross-national, grouped comparison of physical IPV. Measurement-invariance testing over time will inform their utility to monitor SDG5.2.1; cross-national and cross-time measurement-invariance testing of other IPV item sets is warranted.

## Introduction

Intimate partner violence (IPV), defined as psychological, physical, and sexual violence and controlling behaviors perpetrated by a partner, is a significant public health problem. Approximately 27% (95% CI 23%-31%) of ever-partnered women 15-49 years have ever experienced physical and/or sexual IPV, with regional estimates ranging from 18% in Central Asia to 35% in Southern Asia (*1*). Adverse effects of IPV on women include higher rates of economic insecurity and mental-, behavioral-, physical-, sexual-, and reproductive-health conditions among victims than non-victims (*2-8*). IPV compromises national economic development, costing an estimated 5% of world gross domestic product (GDP) and nearly 15% of GDP in Sub-Saharan Africa (*9*).

Given the health, social, and economic costs of IPV, United Nations’ (UN) bodies, treaties, and declarations have called for better statistics on the nature, prevalence, causes, and consequences of violence against women as a basis for its elimination (*10*). This pressure led, in 2015, to Sustainable Development Goal (SDG) 5.2, which urges governments to “eliminate all forms of violence against all women and girls in public and private…” (*11*). Endorsement of SDG5.2 compels national governments to measure and to report rates of violence against women, including IPV (SDG5.2.1).

The decades leading up to SDG5.2 saw marked growth in the number of IPV prevalence surveys, using diverse scales and data-collection approaches (*12*), from small-scale, localized research, to large multi-country studies (*13, 14*), and ongoing surveillance of IPV in multipurpose surveys. No gold standard exists for data collection on IPV, but the Centers for Disease Control and Prevention (CDC) (*15*), World Health Organization (WHO) (*16*), and Demographic and Health Surveys (DHS) (*17*) have agreed best practices. These include direct inquiry within a clear timeframe; use of multiple, behaviorally-specific questions to capture exposure to each type of IPV; reliance on appropriately trained interviewers; and support for respondents and interviewers (*10*).

The DHS domestic violence module (DVM) is the most commonly administered module in lower- and middle-income countries (LMICs) that follows these best practices. The DHS is a flagship project of the United States Agency for International Development (USAID), which has invested several hundred million dollars in data collection since 1984 (*18*) and is a critical source of population and health data for LMICs (*19*). The DHS DVM is optional; however, by the end of 2020, 65 countries had administered it at least once, and 39 countries had administered it more than once (*17*), documenting large differences in national IPV prevalence.(*1*)

While the DHS is used to inform policies, prevention, and response interventions, the DHS DVM has not undergone a rigorous psychometric assessment. It, therefore, is unknown whether questions in the module are measurement invariant across countries on a global scale, a critical precondition for national comparisons. Prior research by this team on DHS questions about the acceptability of IPV found modest non-comparability across settings, due to survey design and contextual factors (*20*). If not identified and accounted for, areas of non-comparability may exaggerate or minimize identified differences in national IPV prevalence (*20*), with potential implications for national policies (*21*). Addressing this knowledge gap now is critical, since the number of countries monitoring IPV will only increase with SDG5.2.

The objective of this paper is to perform the first comprehensive, global psychometric assessment of physical IPV items in the DHS DVM, the most common, standardized module used to measure IPV and controlling behaviors using 36 national surveys in LMICs. We focus our analysis on the seven physical IPV items because they are the most behaviorally specific and are more reliable than the psychological or sexual IPV item sets (*22*). Our findings inform next steps in a global research agenda to improve measures of IPV to monitor SDG5.2. Our use of data from the DHS—the most geographically diverse source for nationally representative data on IPV using identically worded questions—enables us to make evidenced-based recommendations that are global in scope across LMICs.

## Methods

### Eligibility and sample

The DHS is a multipurpose survey administered to large, nationally representative samples of households and randomly selected women of reproductive age (typically 15–49 years) in interviewed households. The DHS routinely collect data on women’s and children’s health. Each country also may include optional modules, like the DVM. The DHS uses standard survey methodology, trained staff, strong mechanisms for quality control, and adherence to internationally recognized guidelines for the ethical collection of data, including on violence against women and girls (VAWG) (*14, 23*).

Eligible countries were those that had completed a DHS between 2012 and 2018 and had administered the same 18 items measuring physical, sexual, or psychological IPV and controlling behaviors. Based on these criteria, the sample for this analysis included 36 DHS conducted in 36 LMICs spanning four continents. Among included countries, response rates at the household level (95.0% to 99.9%), woman level (90.7% to 99.5%), and DV module level (94.2% to 99.9%) were generally high, but these response rates were lower in the Maldives (household 91.6%, woman 84.0%, and DV module 78.0%).

Table 1 summarizes characteristics of the DHS for included countries. Included DHS predominantly represented countries in Sub-Saharan Africa (22 countries), followed by countries in South and Southeast Asia (nine countries), Central Asia (two countries), North Africa/West Asia (two countries), and finally Latin America and the Caribbean (one country). On average, field teams had 6.7 members (range 3.5 to 10.0) and were trained for 29.3 days (range 19.0 to 42.0 days). Most interviews (88.9%) were 30–60 minutes in duration.

**Table 1.**
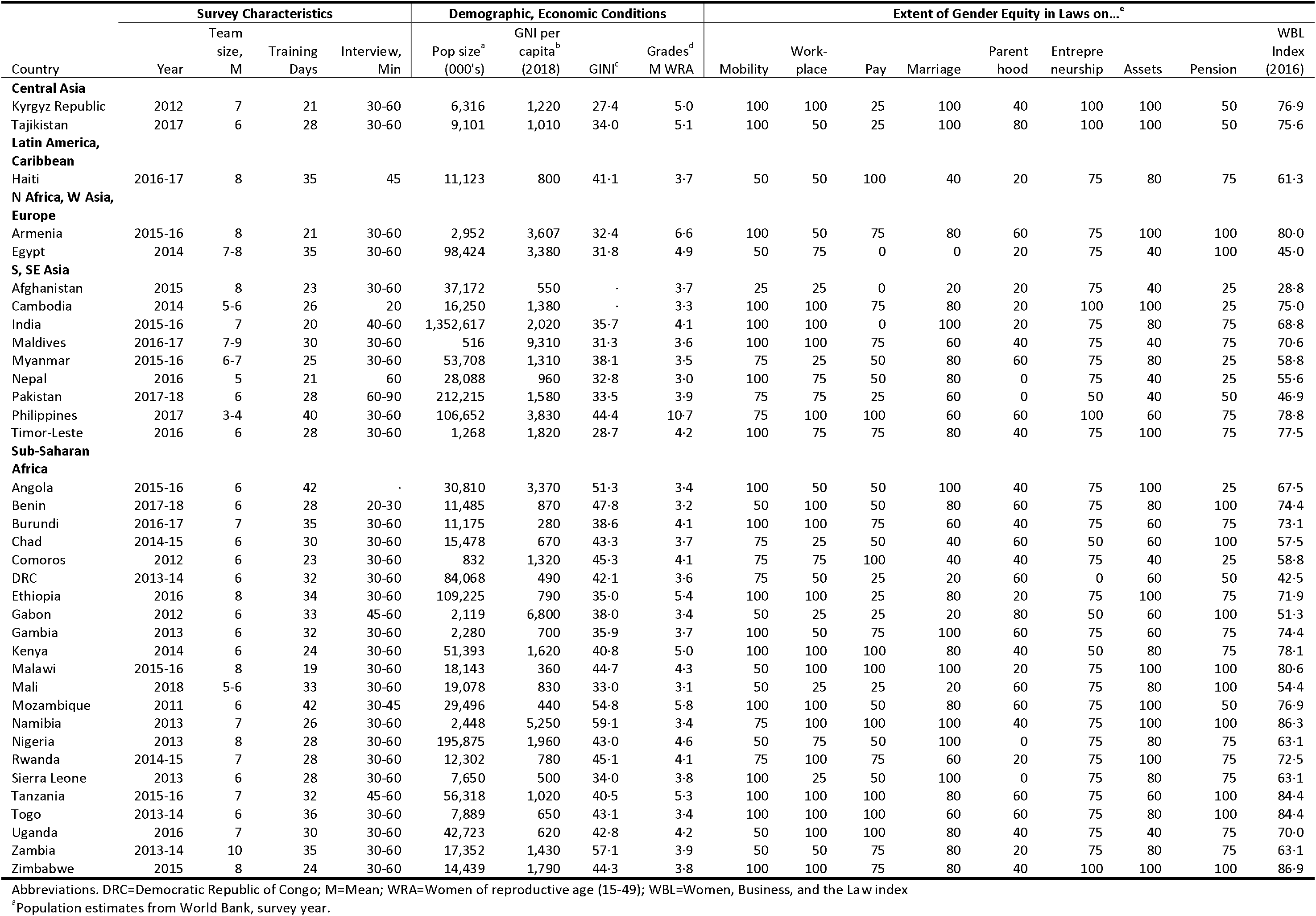
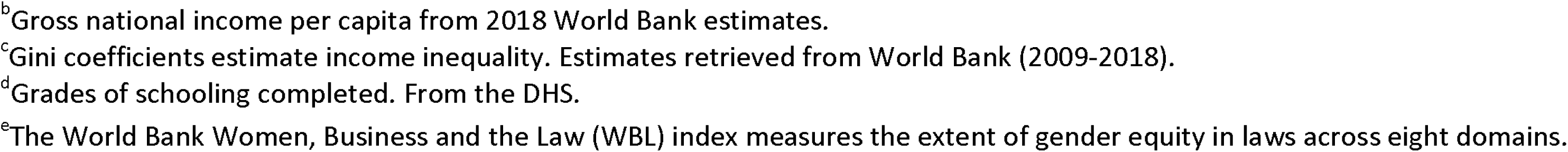
Characteristics of Included Countries and Demographic and Health Surveys, N=36 Surveys across 36 Countries 2012-2018

All included DHS were conducted in LMICs, according to the World Bank classification system. Although a select sample, included DHS were conducted in demographically diverse national populations. For example, countries in the sample ranged widely in population size, from 516,000 people in the Maldives to 1.35 billion in India, and on the GINI index of income inequality, from greater inequality in the Kyrgyz Republic (GINI=27.4) to less inequality in Namibia (GINI=59.1). Countries also ranged in gross national income per capita, from USD280 in Burundi to USD9310 in the Maldives, and in median grades of schooling completed for women of reproductive age (from 3.0 in Nepal to 10.7 in the Philippines). Gender differences in the law, measured using the World Bank index on Women, Business, and the Law, ranged from 28.8 (Afghanistan) to 86.9 (Zimbabwe), with higher scores indicating greater gender parity under the law. (Supplemental Table 1 provides the exact items included in the WBL index.)

### Data on IPV

The IPV-related questions in the DHS DVM (*17*) originated from the Revised Conflict Tactics Scales (*14*), a standardized instrument designed to capture behaviorally based acts of IPV ranging in severity from pushing or shoving to the threat or actual use of a weapon. The DHS DVM has evolved to resemble more closely the instrument used by the WHO (*16*). Specifically, the module includes three items assessing psychological IPV, seven items assessing physical IPV, three items assessing sexual IPV, and five items assessing male controlling behaviors. Occurrence of IPV is measured as the woman’s self-report of exposure to each IPV item: 1) ever in the lifetime of her referent relationship, and if yes, 2) with a standardized frequency in the 12 months before interview. Women’s experience of five controlling behaviors is measured without a specific timeframe or frequency. All items assess IPV in relation to the woman’s most recent spouse or partner. Supplemental Table 2 provides exact item wordings for each IPV item (as translated into English, when the DHS were conducted in other languages). Initial data exploration suggests that fewer than 2% of women in any included DHS sample had missing data on any single IPV item, and 0.02% of women (n=65) or fewer in any included DHS sample had missing data on all IPV items.

**Table 2:** National (Weighted) Estimates for Lifetime and Prior-Year Intimate Partner Violence (IPV), 36 Demographic and Health Surveys across 36 Countries (2012–2018)

|  | Lifetime |  |  |  |  | Prior-Year |  |  |  |  | Cont.<br>Behavior |
| --- | --- | --- | --- | --- | --- | --- | --- | --- | --- | --- | --- |
| Country | Psych. | Phys. | Sexual | Phys. /<br>Sexual | Any | Psych. | Phys. | Sexual | Phys. /<br>Sexual | Any | (any) |
| Central Asia |  |  |  |  |  |  |  |  |  |  |  |
| Kyrgyz Republic | 14.1 | 25.1 | 4.0 | 25.4 | 28.1 | 10.4 | 16.9 | 2.8 | 17.1 | 19.8 | 81.9 |
| Tajikistan | 15.8 | 25.3 | 1.7 | 25.7 | 30.8 | 13.3 | 18.7 | 1.4 | 19.0 | 24.1 | 80.7 |
| Latin America,<br>Caribbean |  |  |  |  |  |  |  |  |  |  |  |
| Haiti | 26.3 | 18.6 | 11.2 | 23.5 | 34.0 | 17.8 | 10.0 | 7.0 | 13.8 | 22.3 | 72.6 |
| N Africa, W Asia,<br>Europe |  |  |  |  |  |  |  |  |  |  |  |
| Armenia | 11.4 | 8.0 | 1.1 | 8.1 | 14.0 | 6.4 | 3.5 | 0.3 | 3.5 | 7.6 | 49.4 |
| Egypt | 18.8 | 25.2 | 4.1 | 25.6 | 30.3 | 13.1 | 13.5 | 2.7 | 14.0 | 18.6 | 78.0 |
| S, SE Asia |  |  |  |  |  |  |  |  |  |  |  |
| Afghanistan | 37.3 | 50.5 | 7.5 | 50.8 | 55.5 | 34.4 | 45.8 | 6.1 | 46.0 | 51.8 | 68.8 |
| Cambodia | 24.8 | 16.2 | 5.5 | 18.2 | 28.7 | 17.3 | 9.3 | 3.9 | 10.9 | 19.6 | 25.9 |
| India | 13.8 | 29.8 | 7.0 | 30.9 | 33.3 | 11.4 | 22.5 | 5.7 | 23.9 | 26.5 | 46.1 |
| Maldives | 11.6 | 12.4 | 2.0 | 12.6 | 17.8 | 7.6 | 5.4 | 0.7 | 5.5 | 10.4 | 38.3 |
| Myanmar | 13.5 | 15.4 | 3.0 | 16.3 | 20.9 | 10.2 | 10.2 | 2.2 | 11.0 | 15.0 | 29.1 |
| Nepal | 12.3 | 22.8 | 7.0 | 24.3 | 26.3 | 7.7 | 10.0 | 4.0 | 11.2 | 13.5 | 34.3 |
| Pakistan | 25.8 | 22.9 | 4.8 | 23.7 | 33.5 | 20.6 | 13.6 | 3.6 | 14.5 | 24.8 | 28.1 |
| Philippines | 10.7 | 11.0 | 4.0 | 12.2 | 16.5 | 6.6 | 4.3 | 2.2 | 5.4 | 9.0 | 37.5 |
| Timor-Leste | 9.4 | 36.6 | 5.0 | 38.1 | 40.1 | 8.9 | 33.1 | 4.8 | 34.6 | 36.8 | 47.3 |
| Sub-Saharan Africa |  |  |  |  |  |  |  |  |  |  |  |
| Angola | 27.7 | 32.5 | 7.7 | 33.9 | 41.3 | 24.0 | 24.2 | 6.7 | 25.8 | 33.8 | 55.9 |
| Benin | 36.7 | 19.5 | 8.8 | 22.4 | 41.8 | 28.7 | 11.1 | 6.1 | 13.9 | 31.8 | 65.3 |
| Burundi | 25.6 | 39.7 | 25.4 | 46.7 | 50.2 | 16.5 | 17.9 | 18.4 | 27.8 | 31.6 | 35.4 |
| Chad | 24.1 | 26.4 | 10.0 | 28.6 | 34.8 | 16.3 | 15.5 | 6.8 | 17.4 | 23.1 | 66.2 |
| Comoros | 8.1 | 5.6 | 1.8 | 6.4 | 10.6 | 6.2 | 4.2 | 1.3 | 4.8 | 8.1 | 66.8 |
| DRC | 36.6 | 45.9 | 25.5 | 50.7 | 57.4 | 29.4 | 30.3 | 19.8 | 36.7 | 43.9 | 82.7 |
| Ethiopia | 24.0 | 23.5 | 10.1 | 26.3 | 33.8 | 20.2 | 16.9 | 8.3 | 19.8 | 27.0 | 56.7 |
| Gabon | 35.1 | 46.2 | 17.0 | 48.6 | 56.1 | 26.6 | 28.3 | 11.8 | 31.2 | 39.2 | 84.7 |
| Gambia | 15.8 | 19.6 | 2.7 | 20.1 | 26.2 | 8.5 | 6.9 | 1.1 | 7.3 | 12.3 | 51.2 |
| Kenya | 32.4 | 36.9 | 13.3 | 39.4 | 47.1 | 23.8 | 22.6 | 9.8 | 25.4 | 32.7 | 63.2 |
| Malawi | 29.5 | 25.9 | 19.2 | 33.8 | 42.2 | 23.0 | 16.2 | 15.4 | 24.1 | 32.6 | 71.4 |
| Mali | 38.4 | 36.8 | 11.8 | 38.5 | 48.9 | 28.1 | 18.0 | 7.8 | 20.9 | 34.0 | 63.7 |
| Mozambique | 14.9 | 18.1 | 3.6 | 18.8 | 23.5 | 12.2 | 14.1 | 2.9 | 14.7 | 18.8 | 39.7 |
| Namibia | 25.0 | 23.4 | 7.6 | 25.0 | 33.3 | 21.0 | 18.7 | 6.6 | 20.2 | 27.8 | 52.3 |
| Nigeria | 19.2 | 14.4 | 4.8 | 16.2 | 24.5 | 15.3 | 9.3 | 3.7 | 11.0 | 19.0 | 63.9 |
| Rwanda | 26.6 | 31.1 | 11.6 | 34.4 | 40.4 | 18.5 | 17.6 | 8.3 | 20.6 | 26.7 | 44.9 |
| Sierra Leone | 29.2 | 44.2 | 7.3 | 45.3 | 50.5 | 20.8 | 27.2 | 5.1 | 28.6 | 33.9 | 79.2 |
| Tanzania | 35.9 | 39.3 | 13.6 | 41.7 | 49.5 | 28.1 | 27.0 | 10.4 | 29.5 | 37.5 | 74.2 |
| Togo | 29.7 | 20.2 | 7.5 | 22.1 | 35.7 | 24.1 | 10.7 | 4.8 | 12.7 | 27.2 | 64.5 |
| Uganda | 41.1 | 40.1 | 22.9 | 46.6 | 55.8 | 29.3 | 22.3 | 16.4 | 29.6 | 39.4 | 71.4 |
| Zambia | 24.0 | 38.8 | 16.7 | 42.7 | 47.1 | 17.8 | 21.3 | 13.0 | 26.5 | 31.1 | 73.8 |
| Zimbabwe | 31.5 | 30.7 | 12.7 | 35.4 | 45.0 | 23.5 | 15.2 | 9.3 | 19.8 | 30.1 | 66.4 |
| Max | 41.1 | 50.5 | 25.5 | 50.8 | 57.4 | 34.4 | 45.8 | 19.8 | 46.0 | 51.8 | 84.7 |
| Min | 8.1 | 5.6 | 1.1 | 6.4 | 10.6 | 6.2 | 3.5 | 0.3 | 3.5 | 7.6 | 25.9 |

Notes. DRC=Democratic Republic of Congo.

### Statistical analysis

The statistical analysis involved four major steps. First, we conducted descriptive analyses to understand country-specific missingness and prevalence for each IPV item and item-specific prevalence ranges across included countries. Given the low estimated prevalences for sexual IPV items, and higher known reliability of behaviorally based physical IPV items, the remaining analyses focused on the seven physical IPV items.

Second, we performed 36 country-specific factor analyses to explore and then to confirm dimensionality of the physical IPV item set, the magnitudes of factor loadings, and overall fit of a unidimensional model. For each country, a unidimensional exploratory factor analysis (EFA) model was considered adequate if all item loadings were 0.35 or greater and if model fit statistics met the following guidelines: the root mean square error of approximation (RMSEA) about 0.08 or lower, and the comparative fit index (CFI) and Tucker-Lewis index (TLI) about 0.95 or higher (*24*). We conducted country-specific confirmatory factor analyses (CFA), including countries that met model-fit criteria in the EFA. We used the same criteria for the item loadings and model fit statistics to assess the adequacy of the fit of all CFA models. The EFA and CFA used the means and variance-adjusted weighted least squares estimators, which are appropriate for dichotomous responses. The approach used pairwise deletion to handle missing data (*25*).

Third, for countries that exhibited adequacy with respect to item loadings and model-fit statistics, we conducted measurement invariance testing, assuming first strict and then approximate measurement invariance. Initially, we performed multiple-group CFA (MGCFA) to test for strict measurement invariance. Because response options for included physical IPV items were dichotomous (1=ever, 0=never experienced the IPV item), we used the Maximum Likelihood estimation to allow us to separately test for metric and scalar invariance (*26*). We assessed configural invariance, or equivalent factor structure; then metric invariance, or equivalent factor loadings; and finally scalar invariance, or equivalent factor loadings and thresholds, across included countries using adjusted likelihood ratio tests.

We used alignment optimization (AO) to assess approximate measurement invariance of the physical IPV items across countries. In the first step (*27*), a model with configural invariance across countries is estimated. In the model, the factor loadings and intercepts are free to vary across countries, whereas the factor means are set equal to zero, and the factor variances fixed at one in all countries. In the second step, the factor means and variances are freed and their values are estimated to minimize the total amount of non-invariance across all parameters. The quality of the alignment result is determined by the percentage of parameters that display non-invariance. As a guide, a limit of 25% of non-invariant parameters or less indicates trustworthy results (*28*). For higher percentages, a Monte Carlo simulation is advised to assess the quality of the results (*28*). Monte Carlo simulations are based on the correlation between the population factor means and the estimated alignment factor means, computed over groups and averaged over replications. Correlations of at least 0.98 produce reliable factor means (*28*). Similar to MGCFA, AO employed maximum likelihood estimation, which uses all available data, assuming data are missing at random (*25, 28*). We used Stata (*29*) for data cleaning and descriptive analyses and Mplus (*30*) for all other analyses.

## Results

### Conventional prevalence estimates of IPV

Estimates for lifetime and prior year IPV were generally high, but ranged widely across sample countries (Table 2). Lifetime psychological IPV ranged from 8.1% in Comoros to 41.1% in Uganda. Lifetime physical IPV ranged from 5.6% in Comoros to 50.5% in Afghanistan. Lifetime sexual IPV ranged from 1.1% in Armenia to 25.5% in the DRC. Combined lifetime physical and/or sexual IPV ranged from 6.4% in Comoros to 50.8% in Afghanistan. Any lifetime IPV ranged from 10.6% in Comoros to 57.4% in the Democratic Republic of Congo (DRC). With respect to prior-year IPV, Armenia had the lowest estimated prevalences for any prior-year IPV (7.6%) and prior-year IPV by most types (physical 3.5%; sexual 0.3%; physical/sexual 3.5%), although prior-year psychological IPV was slightly lowest in Comoros (6.2%). The highest estimated prevalences of prior-year IPV were reported by women in Afghanistan (any 51.8%; psychological 34.4%; physical/sexual 46.0%) and in the Democratic Republic of the Congo (sexual 19.8%). The estimated prevalence of controlling behavior was lowest in Cambodia (25.9%) and highest in Gabon (84.7%).

### Results from country-specific exploratory and confirmatory factor analyses

Table 3 presents results for country-specific EFAs and CFAs for all 36 DHS samples. Across all countries, all loadings exceeded 0.50 in the country-specific EFAs and 0.60 in the country-specific CFAs, above the 0.35 recommended threshold. Moreover, all fit statistics (RMSEA, CFI, TLI) were within recommended thresholds. Thus, based on the country-specific EFAs and CFAs, a unidimensional model for the seven physical IPV items was reasonable across all countries. Loadings for each item and ranges of loadings are reported in Supplemental Table 3.

**Table 3.** Results of Country-Specific Factor Analyses and Alignment Optimization Cross-Country Measurement Invariance Analysis, Seven Physical IPV Items, N=36 Demographic and Health Surveys across 36 Countries (2012-2018)]

| Country | Country-Specific EFAs<sup>1</sup> (N=36) |  |  |  | Country-Specific CFAs<sup>1</sup> (N=36) |  |  |  | Alignment Optimization<sup>2</sup> |
| --- | --- | --- | --- | --- | --- | --- | --- | --- | --- |
|  | Loadings | RMSEA | CFI | TLI | Loadings | RMSEA | CFI | TLI | Non-invariant parameters<br>(intercepts, loadings) |
| **Central Asia** |  |  |  |  |  |  |  |  |  |
| Kyrgyz Republic | 0.84-0.95 | 0.02 | 1.00 | 1.00 | 0.90-0.96 | 0.02 | 1.00 | 1.00 | 2,0 |
| Tajikistan | 0.65-0.95 | 0.02 | 1.00 | 1.00 | 0.74-1.00 | 0.06 | 0.99 | 0.99 | 0,0 |
| **Latin America and the Caribbean** |  |  |  |  |  |  |  |  |  |
| Haiti | 0.70-0.97 | 0.00 | 1.00 | 1.00 | 0.67-0.94 | 0.02 | 1.00 | 1.00 | 3,0 |
| **North Africa, West Asia, Europe** |  |  |  |  |  |  |  |  |  |
| Armenia | 0.95-0.98 | 0.00 | 1.00 | 1.00 | 0.95-1.00 | 0.00 | 1.00 | 1.00 | 3,1 |
| Egypt | 0.83-0.97 | 0.02 | 1.00 | 1.00 | 0.87-0.96 | 0.03 | 1.00 | 1.00 | 2,0 |
| **South and Southeast Asia** |  |  |  |  |  |  |  |  |  |
| Afghanistan | 0.89-0.96 | 0.03 | 0.99 | 0.98 | 0.86-0.97 | 0.03 | 0.99 | 0.98 | 1,0 |
| Cambodia | 0.60-0.94 | 0.02 | 1.00 | 1.00 | 0.82-0.96 | 0.00 | 1.00 | 1.00 | 2,1 |
| India | 0.78-0.94 | 0.02 | 1.00 | 0.99 | 0.81-0.93 | 0.03 | 0.99 | 0.99 | 1,0 |
| Maldives | 0.94-0.98 | 0.01 | 1.00 | 1.00 | 0.73-0.99 | 0.00 | 1.00 | 1.00 | 1,0 |
| Myanmar | 0.78-0.97 | 0.01 | 1.00 | 1.00 | 0.78-0.97 | 0.02 | 1.00 | 1.00 | 2,0 |
| Nepal | 0.84-0.97 | 0.02 | 1.00 | 1.00 | 0.84-0.98 | 0.01 | 1.00 | 1.00 | 1,0 |
| Pakistan | 0.86-0.97 | 0.02 | 1.00 | 1.00 | 0.85-0.98 | 0.05 | 0.99 | 0.99 | 0,1 |
| Philippines | 0.89-0.97 | 0.01 | 1.00 | 1.00 | 0.82-0.95 | 0.01 | 1.00 | 1.00 | 4,1 |
| Timor-Leste | 0.66-0.92 | 0.03 | 0.99 | 0.98 | 0.66-0.93 | 0.03 | 0.99 | 0.99 | 1,0 |
| **Sub-Saharan Africa** |  |  |  |  |  |  |  |  |  |
| Angola | 0.81-0.93 | 0.03 | 1.00 | 0.99 | 0.78-0.94 | 0.02 | 1.00 | 0.99 | 1,0 |
| Benin | 0.81-0.97 | 0.03 | 1.00 | 1.00 | 0.88-0.93 | 0.02 | 1.00 | 0.99 | 1,0 |
| Burundi | 0.84-0.93 | 0.02 | 1.00 | 1.00 | 0.76-0.93 | 0.03 | 1.00 | 0.99 | 2,0 |
| Chad | 0.85-0.95 | 0.04 | 0.99 | 0.99 | 0.82-0.94 | 0.01 | 1.00 | 1.00 | 0,0 |
| Comoros | 0.74-0.99 | 0.02 | 1.00 | 0.99 | 0.82-1.00 | 0.01 | 1.00 | 1.00 | 1,0 |
| DRC | 0.80-0.87 | 0.02 | 0.99 | 0.99 | 0.75-0.91 | 0.03 | 0.99 | 0.98 | 0,0 |
| Ethiopia | 0.76-0.92 | 0.02 | 1.00 | 1.00 | 0.80-0.95 | 0.02 | 1.00 | 0.99 | 0,0 |
| Gabon | 0.64-0.95 | 0.03 | 1.00 | 1.00 | 0.85-0.98 | 0.06 | 0.99 | 0.99 | 3,1 |
| Gambia | 0.57-0.96 | 0.01 | 1.00 | 1.00 | 0.83-1.00 | 0.02 | 0.99 | 0.98 | 2,0 |
| Kenya | 0.79-0.94 | 0.03 | 1.00 | 1.00 | 0.82-0.93 | 0.02 | 1.00 | 1.00 | 1,0 |
| Malawi | 0.83-0.94 | 0.03 | 1.00 | 0.99 | 0.83-0.94 | 0.00 | 1.00 | 1.00 | 3,0 |
| Mali | 0.58-0.91 | 0.01 | 1.00 | 1.00 | 0.74-0.86 | 0.00 | 1.00 | 1.00 | 1,0 |
| Mozambique | 0.83-0.95 | 0.03 | 1.00 | 1.00 | 0.74-0.96 | 0.01 | 1.00 | 1.00 | 2,0 |
| Namibia | 0.87-0.98 | 0.03 | 1.00 | 1.00 | 0.86-0.98 | 0.02 | 1.00 | 1.00 | 1,0 |
| Nigeria | 0.78-0.96 | 0.01 | 1.00 | 1.00 | 0.74-0.96 | 0.02 | 1.00 | 1.00 | 2,1 |
| Rwanda | 0.83-0.95 | 0.02 | 1.00 | 1.00 | 0.84-0.95 | 0.02 | 1.00 | 1.00 | 1,0 |
| Sierra Leone | 0.74-0.95 | 0.03 | 0.99 | 0.99 | 0.74-0.95 | 0.04 | 0.99 | 0.98 | 1,0 |
| Tanzania | 0.76-0.93 | 0.02 | 1.00 | 1.00 | 0.80-0.93 | 0.01 | 1.00 | 1.00 | 2,1 |
| Togo | 0.82-0.95 | 0.02 | 1.00 | 1.00 | 0.85-0.95 | 0.03 | 1.00 | 0.99 | 1,0 |
| Uganda | 0.74-0.93 | 0.02 | 1.00 | 1.00 | 0.75-0.94 | 0.02 | 1.00 | 1.00 | 3,0 |
| Zambia | 0.86-0.91 | 0.02 | 1.00 | 1.00 | 0.79-0.93 | 0.02 | 1.00 | 1.00 | 1,0 |
| Zimbabwe | 0.77-0.93 | 0.02 | 1.00 | 1.00 | 0.79-0.94 | 0.02 | 1.00 | 1.00 | 3,1 |

Notes. DRC=Democratic Republic of Congo.
<sup>1</sup>Model fit criteria (EFA, CFA): root mean square error of approximation (RMSEA) <0.08, comparative fit index (CFI) >0.95, Tucker-Lewis index (TLI) >0.95, loadings >0.35.<sup>2</sup>Alignment optimization model fit criteria: <25% of model estimates non-invariant. Each country has 14 parameter estimates (7 intercepts, 7 loadings).

### Multiple-group CFA and alignment optimization results

The estimation of country-specific unidimensional EFA/CFA models did not require measurement invariance across countries. Table 4 provides the results of the multiple-group CFA, which tests for strict measurement invariance across countries. The metric and configural models differed significantly (*p*<0.001), as did the scalar and metric models (*p*<0.001). Based on the test statistics and their proposed thresholds, strict scalar invariance of the loadings across countries was not achieved.

**Table 4.** Results from Multiple-Group Confirmatory Factor Analysis, N=136,693 across Demographic and Health Surveys in 36 Countries, 2012-2018

| Model | Loglikelihood | Number of parameters | Models compared | Chi-square | Degrees of freedom | *p* value |
| --- | --- | --- | --- | --- | --- | --- |
| Configural | -462468.524 | 539 |  |  |  |  |
| Metric | -463664.754 | 364 | Metric against Configural | 1089.52418 | 175 | <.001 |
| Scalar | -466670.413 | 119 | Scalar against Metric | 4511.56926 | 245 | <.001 |

Considering the MGCFA results, Table 3 includes the results of the alignment optimization approach, which does not assume strict measurement invariance. A guideline of 25% or fewer total non-invariant parameter estimates is recommended for trustworthy latent mean estimates and their comparison across groups. Fifty-five (21.8% of) estimated thresholds, eight (2.8% of) estimated loadings, and 12.3% of all parameter estimates were measurement non-invariant. The items ‘slap’, ‘choke’, and ‘twist’ had a low degree of threshold invariance, and the item ‘choke’ had a low degree of loading invariance.^1^ Overall, these results suggest that the DHS item set for physical IPV exhibits approximate measurement invariance across the 36 countries and allows acceptable alignment performance.

**Table 5.** Results from Alignment Optimization Analysis, N=136,693 across Demographic and Health Surveys in 36 Countries, 2012-2018

| Items | Thresholds |  | Loadings |  |
| --- | --- | --- | --- | --- |
|  | Weighted Avg. Value<br>across Invariant Groups | R<sup>2</sup> | Weighted Avg. Value<br>across Invariant Groups | R<sup>2</sup> |
| Push you, shake you, or throw something at you? | 2.081 | 0.351 | 2.915 | 0.836 |
| Slap you? | 0.263 | 0.000 | 3.867 | 0.394 |
| Punch with his fist or with something that could hurt you? | 3.676 | 0.715 | 3.614 | 0.683 |
| Kick you, drag you, or beat you up? | 3.645 | 0.381 | 3.474 | 0.213 |
| Try to choke you or burn you on purpose? | 5.882 | 0.073 | 2.806 | 0.051 |
| Threaten to attack you with a knife, gun or other weapon? | 6.056 | 0.634 | 2.248 | 0.469 |
| Twist your arm or pull your hair? | 3.220 | 0.000 | 3.599 | 0.359 |

### Model estimates and country rankings on level of physical IPV

Figure 1 provides a visual depiction of country rankings by new and conventional estimates of lifetime physical IPV. Estimated scores derived from the AO approach are provided alongside prevalence estimates with 99.9% confidence intervals using mean estimation with adjustment for sampling. The uncertainties in both sets of estimates account for multiple comparisons. Country rankings, based on both sets of estimates, are provided.^2^ While the distributions of country rankings exhibit some county-level differences, a Wilcoxon matched-pairs sign-rank test supports no significant difference in rankings based on AO and prevalence estimates. Both sets of estimates exhibit a high degree of clustering. For example, in comparing countries using AO-derived scores, 12 clusters emerge wherein country estimates do not differ significantly from one another. In comparing countries by conventional estimates of prevalence and associated confidence limits, three major clusters emerge: countries ranked 1-12, those ranked 13-30, and those ranked 31-36. If we identify clusters across the two distributions in which no country would have a varying rank, we would find that countries ranked 31–36 across both distributions would constitute a “lower physical IPV group” (e.g., lifetime prevalence < 16.0%). Countries ranked 13–30 would constitute an “intermediate physical IPV” group (prevalence 16.0% to < 32.0%), and countries ranked 1–12 would constitute a “higher physical IPV” group (prevalence 32.0% or greater). These cutpoints are provisional and warrant further investigation and possible refinement.

**Figure 1.**
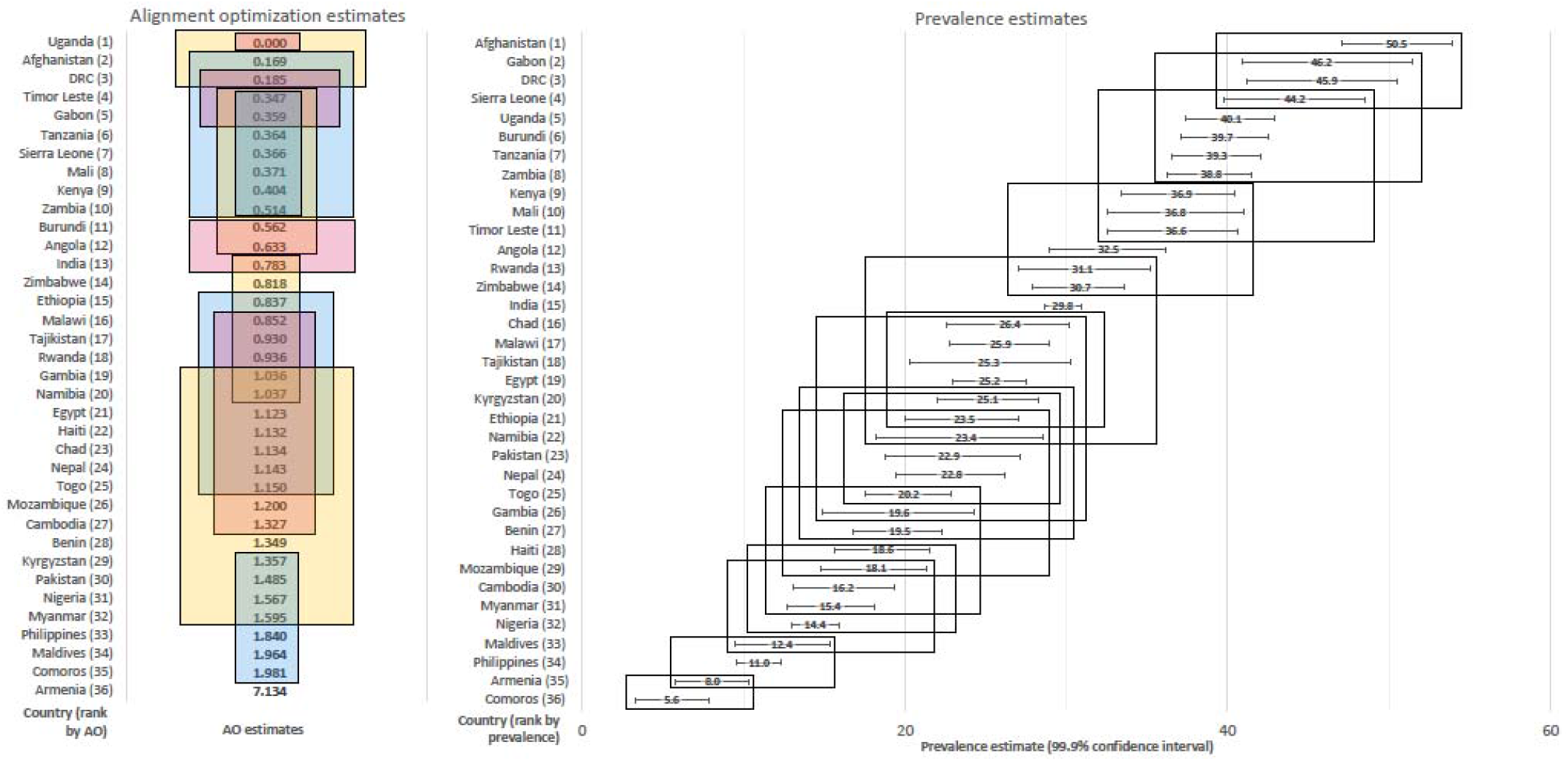
Levels of Physical IPV Derived from the Alignment Optimization Approach and Conventional Prevalence Estimation and Associated Country Rankings, N=36 Demographic and Health Surveys for 36 Countries from 2012-2018

### Convergent Validity of AO-derived Scores and IPV Prevalences

As expected, AO-derived scores were positively correlated (range 0.48-0.66) with prevalence estimates for different types of IPV (Supplemental Figure 1), suggesting convergent validity. Plots provide empirical support for linear relationships.

## Discussion

### Summary of findings

This is the first global analysis of LMICs to assess the measurement invariance of seven standard physical IPV items from the optional domestic violence module administered in 36 Demographic and Health Surveys across 36 LMICs during 2012-2018. Included countries spanned five world regions and had national populations that varied in size, schooling attainment for women, income inequality, and extent of gender equity of the legal environment. This item set exhibited good country-specific measurement properties for all 36 LMICs. While this item set did not meet the criteria for strict measurement invariance across countries, it did meet the criteria for approximate invariance across all 36 LMICs. The distributions of country rankings, based on alignment-optimization derived scores and the more standard lifetime estimated prevalences for physical IPV, were similar. However, AO-derived scores and prevalences for lifetime physical IPV often were highly clustered and not significantly different, suggesting that individual country rankings were not interpretable using either set of estimates. Instead, we recommend making grouped country comparisons according to ranges in the level of physical IPV. Given the priority of SDG5.2 to eliminate all violence against women, we suggest groupings that distinguish the primary goal of “elimination” (0%) from the clustered ranges of 1% to <16%, 16.0% to <32.0%, and 32.0% or higher.

### Limitations and strengths

Findings from this analysis are limited to the seven included physical IPV items and cannot be extrapolated to different physical IPV items, or other forms of IPV. Findings also are limited to this non-representative set of LMICs and for the period of analysis (2012-2018). That said, demonstrating approximate non-invariance of a substantial set of physical IPV items across highly diverse LMICs spanning five regions strongly suggests the utility of this item set to compare groups of countries on behaviorally specific measures of physical IPV. These results support their use to monitor SDG5.2.1 for physical IPV.

### Implications for research and policy

Implications for research and the global monitoring of IPV are notable. First, replication of this study in higher-income countries and under-represented LMIC-regions is needed. Second, the proposed ranges to distinguish groups of countries on levels of physical IPV should be considered a guide that addresses both the high clustering of countries on estimates and the ultimate goal of eliminating all violence against women. More research is needed to assess whether the proposed ranges to identify groups of countries on levels of physical IPV are useful worldwide. Third, cognitive testing of ‘slap’, ‘twist’, and ‘choke’ items is needed to improve cross-national measurement equivalence of these items and the overall scale. We recommend this approach over dropping or separating these items into another scale based on both the evidence presented here that the seven items are unidimensional as well as the importance of preserving content validity and reducing under-estimation for levels of physical IPV. Fourth, further testing of the item set for measurement invariance across repeated national surveys is needed to assess how invariant these physical IPV items are over extended periods of time. Fifth, this analysis should be replicated for the controlling behaviors and expanded item sets to measure psychological IPV and sexual IPV (currently only three items each). Until then, the seven physical IPV items from the DHS DVM appear useful to compare country groupings across clustered levels of national physical IPV, as a general guide toward the ultimate SDG5.2 goal of elimination.

## Conclusion

Alignment optimization is a powerful approach to assess approximate measurement equivalence of scales across countries charged with monitoring the SDGs. The seven physical IPV items from the DHS DV module exhibit approximate measurement invariance across 36 diverse LMICs, and if shown to be time-invariant and invariant across HICs, may be useful to monitor SDG5.2 globally.

## Data Availability

Data from the Demographic and Health Surveys (DHS) are publicly available upon reasonable request to Measure DHS: https://dhsprogram.com/data/new-user-registration.cfm. Investigators must request from Measure DHS access to the data used in this analysis.

## Acknowledgements

The authors thank our advisory board (Kristin Dunkle, Claudia Garcia-Moreno, Enrique Gracia, Andrew Gibbs, Sunita Kishor, Rachel Jewkes) for comments on the analysis and interpretation.

## Funding

*Eunice Kennedy Shriver* National Institute of Child Health and Human Development and the National Institute of Mental Health R01HD099224 (PIs CJC, KMY).

## Author contributions

Conceptualization: KMY, YFC, CJC

Methodology: KMY, YFC, CJC

Investigation: KMY, YFC, ZK

Visualization: KMY, YFC, ZK

Funding Acquisition: KMY, CJC

Project administration: KMY, CJC, IB, ZK

Supervision: KMY, YFC, CJC, NK

Writing – original draft: KMY, YFC, ZK, IB, CJC

Writing – reviewing and editing: KMY, YFC, ZK, IB, CJC, NK

## Competing interests

Authors declare that they have no competing interests.

## Supplementary material

**Table S1**. Items Included in the World Bank’s Women, Business and the Law Index (Women, Business, and Law Data for 2016)

**Table S2**. Items included in the DHS Domestic Violence module

**Table S3**. Item Loadings from Exploratory and Confirmatory Factor Analyses of Physical IPV, N=36 Demographic and Health Surveys across 36 Countries (2012-2018)

**Figure S1**. Correlations of Prevalence Estimates and Alignment Optimization Estimates of Lifetime Intimate Partner Violence across 36 Countries, 2012-2018.

## Footnotes

1 As the bivariate tables of the indicators had empty cells in many countries, not all the variables could be used to perform the simulation. Their presence also led to problems with the estimates of the residual covariance matrix. We thus relied on the 25% criterion.

2 The prevalence estimates are based on the aggregates of the observed responses to intimate partner violence indicators. The factor means of the AO approach are based on a factor model that presumes that observed indicators reflect a latent IPV construct. The endorsement of an indicator is a function of a threshold parameter, which can be conceptualized as a cutoff score at which a woman participant transitions from the report of none to the presence of an act, and a factor loading that relates the variable to the factor.

## Notes

### Competing Interest Statement

The authors have declared no competing interest.

### Author Declarations

Emory University determined the study to be exempt from IRB review, as the data analyzed in the study are fully de-identified, publicly available datasets and no one involved in the original studies is engaged in the research.

## References

1. U. World Health Organization on behalf of the United Nations Inter-Agency Working Group on Violence Against Women Estimation and Data (UNICEF, UNODC, UNSD, UNWomen), “Violence against women prevalence estimates 2018: Global, regional and national prevalence estimates for intimate partner violence against women and global and regional prevalence estimates for non-partner sexual violence against women,” (World Health Organization, Geneva, 2021).

2. K. M. Devries et al., Intimate partner violence and incident depressive symptoms and suicide attempts: a systematic review of longitudinal studies. PLoS Med 10, e1001439 (2013).

3. K. M. Devries et al., Intimate partner violence victimization and alcohol consumption in women: a systematic review and meta-analysis. Addiction 109, 379–391 (2014).

4. G. Dillon, R. Hussain, D. Loxton, S. Rahman, Mental and Physical Health and Intimate Partner Violence against Women: A Review of the Literature. Int J Family Med 2013, 313909 (2013).

5. C. A. Crane, S. W. Hawes, A. H. Weinberger, Intimate partner violence victimization and cigarette smoking: a meta-analytic review. Trauma, violence & abuse 14, 305–315 (2013).

6. H. A. Beydoun, M. A. Beydoun, J. S. Kaufman, B. Lo, A. B. Zonderman, Intimate partner violence against adult women and its association with major depressive disorder, depressive symptoms and postpartum depression: a systematic review and meta-analysis. Soc Sci Med 75, 959–975 (2012).

7. L. Maxwell, K. Devries, D. Zionts, J. L. Alhusen, J. Campbell, Estimating the effect of intimate partner violence on women’s use of contraception: a systematic review and meta-analysis. PLoS One 10, e0118234 (2015).

8. K. M. Yount, Resources, family organization, and domestic violence against married women in Minya, Egypt. Journal of Marriage & Family 67, 579–596 (2005).

9. A. Hoeffler, J. Fearon, “Benefits and costs of the conflict and violence targets for the post-2015 development agenda,” Post-2015 Consensus, Conflict and Violence Assessment Paper (Copenhagen Consensus Center, Copenhagen, 2014).

10. United Nations Department of Economic and Social Affairs Statistics Division, “Guidelines for producing statistics on violence against women-statistical surveys,” (United Nations New York, 2014).

11. Transforming Our World: the 2030 Agenda for Sustainable Development (2015).

12. K. M. Devries et al., Global health. The global prevalence of intimate partner violence against women. Science 340, 1527–1528 (2013).

13. C. Garcia-Moreno, H. A. Jansen, M. Ellsberg, L. Heise, C. H. Watts, Prevalence of intimate partner violence: findings from the WHO multi-country study on women’s health and domestic violence. Lancet 368, 1260–1269 (2006).

14. S. Kishor, K. Johnson, Profiling Domestic Violence: A Multi-Country Study. (ORC Macro, Calverton, Maryland., 2004).

15. M. J. Breiding, K. C. Basile, S. G. Smith, M. C. Black, R. R. Mahendra, “Intimate partner violence surveillance: uniform definitions and recommended data elements, version 2.0,” (National Center for Injury Prevention and Control, Centers for Disease Control and Prevention, Atlanta, GA, 2015).

16. World Health Organization, “WHO multi-country study on women’s health and domestic violence against women: summary report of initial results onprevalence, health outcomes and women’s responses,” (World Health Organization, Geneva, 2005).

17. MEASURE DHS, ICF International, “Domestic Violence Module: Demographic and Health Surveys Methodology,” (MEASURE DHS/ICF International, Calverton, MD, 2014).

18. M. Short Fabic, Y. Choi, S. Bird, A systematic review of Demographic and Health Surveys: data availability and utilization for research. Bull World Health Organ 90, 604–612 (2012).

19. A. Hancioglu, F. Arnold, Measuring coverage in MNCH: tracking progress in health for women and children using DHS and MICS household surveys. PLoS Med 10, e1001391 (2013).

20. K. M. Yount, N. Halim, M. Hynes, E. R. Hillman, Response effects to attitudinal questions about domestic violence against women: a comparative perspective. Social Science Research 40, 873–884 (2011).

21. N. Guenole, A. Brown, The consequences of ignoring measurement invariance for path coefficients in structural equation models. Front Psychol 5, 980 (2014).

22. D. Costa, H. Barros, Instruments to assess intimate partner violence: A scoping review of the literature. Violence and victims 31, 591–621 (2016).

23. M. Ellsberg, L. Heise, Bearing witness: ethics in domestic violence research. Lancet 359, 1599–1604 (2002).

24. L. Hu, P. M. Bentler, Cutoff criteria for fit indexes in covariance structure analysis: Conventional criteria versus new alternatives. Structural Equation Modeling: A Multidisciplinary Journal 6, 1–55 (1999).

25. T. A. Brown, Confirmatory factor analysis for applied research. (The Guilford Press, London, 2006).

26. B. Muthén, T. Asparouhov, IRT studies of many groups: the alignment method. Frontiers in Psychology 5, 1–7 (2014).

27. B. Muthén, T. Asparouhov, Recent methods for the study of measurement invariance with many groups: Alignment and random effects. Sociological Methods & Research 47, 637–664 (2018).

28. T. Asparouhov, B. Muthén, Multiple-group factor analysis alignment. Structural Equation Modeling: A Multidisciplinary Journal 21, 495–508 (2014).

29. StataCorp, Stata statisical software: Release 16. (StatCorp LP., College Station, TX, 2019).

30. L. K. Muthén, B. O. Muthén, Mplus user’s guide. null (Muthén & Muthén, Los Angeles: CA, ed. 8th, 1998-2018).

