## Supplemental Files for "Global Measurement of Physical Intimate Partner Violence to Monitor Sustainable Development Goal 5"

**Affiliations:**

Hubert Department of Global Health

Rollins School of Public Health

Emory University

1518 Clifton Rd NE, Atlanta, GA 30322

**Abstract**

**Background.** One third of women experience IPV and potential sequelae. Sustainable Development Goal (SDG) 5.2—to eliminate all violence against women, including IPV—compels national governments to monitor such violence. We conducted the first global measurement-invariance assessment of standardized physical IPV items.

**Methods.** Thirty-six Demographic and Health Surveys (DHS) from 36 Lower-/Middle-Income Countries (LMICs) administering the same 18 IPV items during 2012-2018 were included. We performed exploratory and confirmatory factor analyses (EFA/CFA) with seven physical IPV items, which are the most behaviorally specific and reliable. Datasets meeting EFA/CFA model fit criteria (loadings>.35, RMSEA<.08, CFI/TLI>.95) were included in multiple-group CFA to test strict measurement invariance, and in alignment optimization (AO) to test approximate measurement invariance. We compared national rankings based on AO-derived scores and lifetime physical IPV prevalences, and correlated AO-dervied scores with physical, sexual, and psychological IPV prevalences.

**Results.** Estimated lifetime physical IPV varied widely (5.6%-50.5%). All loadings and fit statistics met thresholds in country-specific EFA/CFAs. A unidimensional, seven-item physical IPV construct lacked scalar invariance in multiple-group CFA but achieved approximate measurement invariance in AO analysis, as 12.3% (<25%) of model parameters were non-invariant. National rankings of AO-derived scores and estimated physical IPV prevalences were similarly distributed, but national estimates often were not significantly different, so grouped score ranges or prevalence ranges are advised. Three items (slap, twist, choke) warrant cognitive testing to improve their psychometric performance. Correlations of AO-derived scores with IPV prevalences ranged from .48 to .66.

**Conclusions.** Seven DHS physical-IPV items were approximately invariant across 36 LMICs spanning five regions and are reasonable for cross-national, grouped comparison of physical IPV. Measurement-invariance testing over time will inform their utility to monitor SDG5.2.1; cross-national and cross-time measurement-invariance testing of other IPV item sets is warranted.

**This file includes:**

**Table S1.** Items Included in the World Bank's Women, Business and the Law Index (Women, Business, and Law Data for 2016)

**Table S2.** Items included in the DHS Domestic Violence module

**Table S3.** Item Loadings from Exploratory and Confirmatory Factor Analyses of Physical IPV, N=36 Demographic and Health Surveys across 36 Countries (2012-2018)

**Figure S1.** Correlations of Prevalence Estimates and Alignment Optimization Estimates of Lifetime Intimate Partner Violence across 35 Countries (outlier removed), 2012-2018.

**Table S1.** Items Included in the World Bank's Women, Business and the Law Index (Women, Business, and Law Data for 2016)

| **Country** | WBL Index | **Mobility** | | | | **Workplace** | | | | **Pay** | | |  |  |
| --- | --- | --- | --- | --- | --- | --- | --- | --- | --- | --- | --- | --- | --- | --- |
|  |  | Apply for passport | Travel outside country | Travel outside her home | Choose where to live | Get a job | Law prohibits gender discrimination in employment | Legislation on sexual harassment in employment | Criminal penalties or civil remedies for workplace sexual harassment | Equal renumeration for work of equal value | Same night hours | Work in jobs deemed dangerous | | Work in the same industries as men |
| **Central Asia** |  |  |  |  |  |  |  |  |  |  |  |  | |  |
| Kyrgyz Republic | 76.9 | 1 | 1 | 1 | 1 | 1 | 1 | 1 | 1 | 0 | 1 | 0 | | 0 |
| Tajikistan | 75.6 | 1 | 1 | 1 | 1 | 1 | 1 | 0 | 0 | 1 | 0 | 0 | | 0 |
| Haiti | 61.3 | 1 | 1 | 1 | 1 | 1 | 1 | 1 | 1 | 1 | 1 | 1 | | 1 |
| Armenia | 80.0 | 1 | 1 | 1 | 1 | 1 | 1 | 0 | 0 | 0 | 1 | 1 | | 1 |
| Egypt | 45.0 | 0 | 1 | 0 | 1 | 0 | 1 | 1 | 1 | 0 | 0 | 0 | | 0 |
| Afghanistan | 28.8 | 0 | 1 | 0 | 0 | 1 | 0 | 0 | 0 | 0 | 0 | 0 | | 0 |
| Cambodia | 75.0 | 1 | 1 | 1 | 1 | 1 | 1 | 1 | 1 | 0 | 1 | 1 | | 1 |
| India | 68.8 | 1 | 1 | 1 | 1 | 1 | 1 | 1 | 1 | 0 | 0 | 0 | | 0 |
| Maldives | 70.6 | 1 | 1 | 1 | 1 | 1 | 1 | 1 | 1 | 0 | 1 | 1 | | 1 |
| Myanmar | 58.8 | 0 | 1 | 1 | 1 | 1 | 0 | 0 | 0 | 0 | 1 | 1 | | 0 |
| Nepal | 55.6 | 1 | 1 | 1 | 1 | 1 | 0 | 1 | 1 | 0 | 0 | 1 | | 1 |
| Pakistan | 46.9 | 0 | 1 | 1 | 1 | 1 | 0 | 1 | 1 | 0 | 0 | 1 | | 0 |
| Philippines | 78.8 | 0 | 1 | 1 | 1 | 1 | 1 | 1 | 1 | 1 | 1 | 1 | | 1 |
| Timor-Leste | 77.5 | 1 | 1 | 1 | 1 | 1 | 1 | 1 | 0 | 0 | 1 | 1 | | 1 |
| Angola | 67.5 | 1 | 1 | 1 | 1 | 1 | 1 | 0 | 0 | 1 | 1 | 0 | | 0 |
| Benin | 74.4 | 0 | 1 | 1 | 0 | 1 | 1 | 1 | 1 | 1 | 1 | 0 | | 0 |
| Burundi | 73.1 | 1 | 1 | 1 | 1 | 1 | 1 | 1 | 1 | 0 | 1 | 1 | | 1 |
| Chad | 57.5 | 1 | 1 | 1 | 0 | 0 | 1 | 0 | 0 | 1 | 1 | 0 | | 0 |
| Comoros | 58.8 | 1 | 1 | 1 | 0 | 0 | 1 | 1 | 1 | 1 | 1 | 1 | | 1 |
| DRC | 42.5 | 1 | 1 | 1 | 0 | 0 | 0 | 1 | 1 | 0 | 1 | 0 | | 0 |
| Ethiopia | 71.9 | 1 | 1 | 1 | 1 | 1 | 1 | 1 | 1 | 0 | 1 | 0 | | 0 |
| Gabon | 51.3 | 0 | 1 | 1 | 0 | 0 | 1 | 0 | 0 | 0 | 1 | 0 | | 0 |
| Gambia | 74.4 | 1 | 1 | 1 | 1 | 1 | 1 | 0 | 0 | 0 | 1 | 1 | | 1 |
| Kenya | 78.1 | 1 | 1 | 1 | 1 | 1 | 1 | 1 | 1 | 1 | 1 | 1 | | 1 |
| Malawi | 80.6 | 0 | 1 | 1 | 0 | 1 | 1 | 1 | 1 | 1 | 1 | 1 | | 1 |
| Mali | 54.4 | 0 | 1 | 1 | 0 | 1 | 0 | 0 | 0 | 0 | 1 | 0 | | 0 |
| Mozambique | 76.9 | 1 | 1 | 1 | 1 | 1 | 1 | 1 | 1 | 0 | 1 | 0 | | 1 |
| Namibia | 86.3 | 0 | 1 | 1 | 1 | 1 | 1 | 1 | 1 | 1 | 1 | 1 | | 1 |
| Nigeria | 63.1 | 0 | 1 | 1 | 0 | 1 | 0 | 1 | 1 | 0 | 1 | 1 | | 0 |
| Rwanda | 72.5 | 1 | 1 | 1 | 0 | 1 | 1 | 1 | 1 | 0 | 1 | 1 | | 1 |
| Sierra Leone | 63.1 | 1 | 1 | 1 | 1 | 1 | 0 | 0 | 0 | 0 | 1 | 1 | | 0 |
| Tanzania | 84.4 | 1 | 1 | 1 | 1 | 1 | 1 | 1 | 1 | 1 | 1 | 1 | | 1 |
| Togo | 84.4 | 1 | 1 | 1 | 1 | 1 | 1 | 1 | 1 | 1 | 1 | 1 | | 1 |
| Uganda | 70.0 | 0 | 1 | 1 | 0 | 1 | 1 | 1 | 1 | 1 | 1 | 1 | | 1 |
| Zambia | 63.125 | 0 | 1 | 1 | 0 | 1 | 1 | 0 | 0 | 0 | 1 | 1 | | 1 |
| Zimbabwe | 86.9 | 1 | 1 | 1 | 1 | 1 | 1 | 1 | 1 | 0 | 1 | 1 | | 1 |

| **Marriage** | | | | | **Parenthood** | | | | | **Entrepreneurship** | | | |
| --- | --- | --- | --- | --- | --- | --- | --- | --- | --- | --- | --- | --- | --- |
| No legal provision to obey husband | Woman can be head of household or family | Domestic violence legislation | Obtain divorce | Right to remarry | At least 14 weeks paid leave | Government administers 100% of maternity leave benefits | Paid paternal leave | Paid parental leave | Dismissal of pregant workers prohibited | Sign contract | Register business | Open bank account | Prohibit gender discrimination in access to credit |
| 1 | 1 | 1 | 1 | 1 | 1 | 0 | 0 | 0 | 1 | 1 | 1 | 1 | 1 |
| 1 | 1 | 1 | 1 | 1 | 1 | 1 | 0 | 1 | 1 | 1 | 1 | 1 | 1 |
| 1 | 1 | 0 | 1 | 1 | 0 | 0 | 1 | 0 | 1 | 1 | 0 | 1 | 0 |
| 1 | 1 | 0 | 1 | 1 | 1 | 1 | 0 | 1 | 1 | 1 | 1 | 1 | 0 |
| 0 | 0 | 0 | 0 | 0 | 0 | 0 | 0 | 0 | 1 | 1 | 1 | 1 | 0 |
| 0 | 1 | 0 | 0 | 0 | 0 | 0 | 1 | 0 | 0 | 1 | 1 | 1 | 0 |
| 1 | 1 | 1 | 1 | 0 | 0 | 0 | 0 | 0 | 1 | 1 | 1 | 1 | 1 |
| 1 | 1 | 1 | 1 | 1 | 0 | 0 | 0 | 0 | 1 | 1 | 1 | 1 | 0 |
| 1 | 1 | 1 | 0 | 0 | 0 | 0 | 1 | 0 | 1 | 1 | 1 | 1 | 0 |
| 1 | 1 | 0 | 1 | 1 | 1 | 1 | 1 | 0 | 0 | 1 | 1 | 1 | 0 |
| 1 | 1 | 1 | 0 | 1 | 0 | 0 | 0 | 0 | 0 | 1 | 1 | 1 | 0 |
| 1 | 1 | 1 | 0 | 0 | 0 | 0 | 0 | 0 | 0 | 1 | 0 | 1 | 0 |
| 1 | 1 | 1 | 0 | 0 | 0 | 1 | 1 | 0 | 1 | 1 | 1 | 1 | 1 |
| 1 | 1 | 1 | 1 | 0 | 0 | 0 | 1 | 0 | 1 | 1 | 1 | 1 | 0 |
| 1 | 1 | 1 | 1 | 1 | 0 | 1 | 0 | 0 | 1 | 1 | 1 | 1 | 0 |
| 1 | 1 | 1 | 1 | 0 | 1 | 0 | 1 | 0 | 1 | 1 | 1 | 1 | 0 |
| 1 | 0 | 1 | 1 | 0 | 0 | 0 | 1 | 0 | 1 | 1 | 1 | 1 | 0 |
| 1 | 0 | 0 | 1 | 0 | 1 | 0 | 1 | 0 | 1 | 1 | 1 | 0 | 0 |
| 1 | 0 | 1 | 0 | 0 | 1 | 0 | 0 | 0 | 1 | 1 | 1 | 1 | 0 |
| 0 | 0 | 0 | 1 | 0 | 1 | 0 | 1 | 0 | 1 | 0 | 0 | 0 | 0 |
| 1 | 1 | 1 | 1 | 0 | 0 | 0 | 0 | 0 | 1 | 1 | 1 | 1 | 0 |
| 0 | 0 | 0 | 1 | 0 | 1 | 1 | 1 | 0 | 1 | 1 | 1 | 0 | 0 |
| 1 | 1 | 1 | 1 | 1 | 1 | 0 | 1 | 0 | 1 | 1 | 1 | 1 | 0 |
| 1 | 1 | 0 | 1 | 1 | 0 | 0 | 1 | 0 | 1 | 1 | 0 | 1 | 0 |
| 1 | 1 | 1 | 1 | 1 | 0 | 0 | 0 | 0 | 1 | 1 | 1 | 1 | 0 |
| 0 | 0 | 0 | 1 | 0 | 1 | 1 | 1 | 0 | 0 | 1 | 1 | 1 | 0 |
| 1 | 1 | 1 | 1 | 0 | 0 | 1 | 1 | 0 | 1 | 1 | 1 | 1 | 0 |
| 1 | 1 | 1 | 1 | 1 | 0 | 1 | 0 | 0 | 1 | 1 | 1 | 1 | 0 |
| 1 | 1 | 1 | 1 | 1 | 0 | 0 | 0 | 0 | 0 | 1 | 1 | 1 | 0 |
| 1 | 0 | 1 | 1 | 0 | 0 | 0 | 1 | 0 | 0 | 1 | 1 | 1 | 0 |
| 1 | 1 | 1 | 1 | 1 | 0 | 0 | 0 | 0 | 0 | 1 | 1 | 1 | 0 |
| 1 | 1 | 0 | 1 | 1 | 0 | 1 | 1 | 0 | 1 | 1 | 1 | 1 | 0 |
| 1 | 1 | 0 | 1 | 0 | 1 | 0 | 1 | 0 | 1 | 1 | 1 | 1 | 0 |
| 1 | 1 | 1 | 0 | 1 | 0 | 0 | 1 | 0 | 1 | 1 | 1 | 1 | 0 |
| 1 | 1 | 1 | 0 | 1 | 0 | 0 | 0 | 0 | 1 | 1 | 1 | 1 | 0 |
| 1 | 1 | 1 | 0 | 1 | 1 | 0 | 0 | 0 | 1 | 1 | 1 | 1 | 1 |

| **Assets** | | | | | **Pension** | | | |
| --- | --- | --- | --- | --- | --- | --- | --- | --- |
| Equal property ownership | Equal inheritance rights (sons and daughters) | Equal spousal inheritance rights | Equal administrative authority over assets | Valuation of nonmonetary contributions | Full pension benefits | Partial pension benefits | Mandatory retirement age | Periods of absence due to child care accounted for in pension |
| 1 | 1 | 1 | 1 | 1 | 0 | 0 | 1 | 1 |
| 1 | 1 | 1 | 1 | 1 | 0 | 0 | 1 | 1 |
| 1 | 1 | 0 | 1 | 1 | 1 | 1 | 1 | 0 |
| 1 | 1 | 1 | 1 | 1 | 1 | 1 | 1 | 1 |
| 1 | 0 | 0 | 1 | 0 | 1 | 1 | 1 | 1 |
| 1 | 0 | 0 | 1 | 0 | 0 | 0 | 1 | 0 |
| 1 | 1 | 1 | 1 | 1 | 0 | 0 | 1 | 0 |
| 1 | 1 | 1 | 1 | 0 | 1 | 1 | 1 | 0 |
| 1 | 0 | 0 | 1 | 0 | 1 | 1 | 1 | 0 |
| 1 | 1 | 1 | 1 | 0 | 0 | 0 | 1 | 0 |
| 1 | 0 | 0 | 1 | 0 | 0 | 0 | 1 | 0 |
| 1 | 0 | 0 | 1 | 0 | 0 | 0 | 1 | 1 |
| 0 | 1 | 1 | 0 | 1 | 1 | 1 | 1 | 0 |
| 1 | 1 | 1 | 1 | 1 | 1 | 1 | 1 | 0 |
| 1 | 1 | 1 | 1 | 1 | 0 | 1 | 0 | 0 |
| 1 | 1 | 1 | 1 | 0 | 1 | 1 | 1 | 1 |
| 1 | 0 | 0 | 1 | 1 | 1 | 1 | 1 | 0 |
| 0 | 1 | 1 | 0 | 1 | 1 | 1 | 1 | 1 |
| 1 | 0 | 0 | 1 | 0 | 0 | 0 | 1 | 0 |
| 0 | 1 | 1 | 0 | 1 | 0 | 1 | 1 | 0 |
| 1 | 1 | 1 | 1 | 1 | 1 | 1 | 1 | 0 |
| 0 | 1 | 1 | 0 | 1 | 1 | 1 | 1 | 1 |
| 1 | 0 | 0 | 1 | 1 | 1 | 1 | 1 | 0 |
| 1 | 1 | 0 | 1 | 1 | 1 | 1 | 1 | 0 |
| 1 | 1 | 1 | 1 | 1 | 1 | 1 | 1 | 1 |
| 1 | 1 | 1 | 1 | 0 | 1 | 1 | 1 | 1 |
| 1 | 1 | 1 | 1 | 1 | 0 | 1 | 0 | 1 |
| 1 | 1 | 1 | 1 | 1 | 1 | 1 | 1 | 1 |
| 1 | 1 | 1 | 1 | 0 | 1 | 1 | 1 | 0 |
| 1 | 1 | 1 | 1 | 1 | 1 | 1 | 1 | 0 |
| 1 | 1 | 1 | 1 | 0 | 1 | 1 | 1 | 0 |
| 1 | 0 | 0 | 1 | 1 | 1 | 1 | 1 | 1 |
| 1 | 1 | 1 | 1 | 0 | 1 | 1 | 1 | 1 |
| 1 | 0 | 0 | 1 | 0 | 1 | 1 | 1 | 0 |
| 1 | 1 | 1 | 1 | 0 | 1 | 1 | 1 | 0 |
| 1 | 1 | 1 | 1 | 1 | 1 | 1 | 1 | 1 |

Notes. DRC=Democratic Republic of Congo; Figures for the GINI are for the years 2009-2018.

^a^Subnational.

**Table S2.** Items included in the DHS Domestic Violence module

| **Supplemental Table 2. Items included in the DHS Domestic Violence module** | |
| --- | --- |
| **Controlling behaviors** | |
| *First, I am going to ask you about some situations which happen to some women. Please tell me if these apply to your relationship with your (last) (husband/partner)?* | |
| *(Response options: Yes, No, Don't Know)* | |
| a | He (is/was) jealous or angry if you (talk/talked) to other men? |
| b | He frequently (accuses/accused) you of being unfaithful? |
| c | He (does/did) not permit you to meet your female friends? |
| d | He (tries/tried) to limit your contact with your family? |
| e | He (insists/insisted) on knowing where you (are/were) at all times? |
| **Emotional IPV** | |
| *Now I need to ask some more questions about your relationship with your (last) (husband/partner).* | |
| *(Response options: Yes, No)* | |
| a | say or do something to humiliate you in front of others? |
| b | threaten to hurt or harm you or someone you care about? |
| c | insult you or make you feel bad about yourself? |
| **Physical IPV** | |
| *Did your (last) (husband/partner) ever do any of the following things to you:* | |
| *(Response options: Yes, No)* | |
| a | Push you, shake you, or throw something at you? |
| b | Slap you? |
| c | Twist your arm or pull your hair? |
| d | Punch with his fist or with something that could hurt you? |
| e | Kick you, drag you, or beat you up? |
| f | Try to choke you or burn you on purpose? |
| g | Threaten to attack you with a knife, gun or other weapon? |
| **Sexual IPV** | |
| *Did your (last) (husband/partner) ever do any of the following things to you:* | |
| *(Response options: Yes, No)* | |
| h | Physically force you to have sexual intercourse with him even when you did not want to? |
| i | Physically force you to perform any other sexual acts you did not want to? |
| j | Force you with threats or any other way to perform sexual acts that you did not want to do? |

**Table S3.** Item Loadings from Exploratory and Confirmatory Factor Analyses of Physical IPV, N=36 Demographic and Health Surveys across 36 Countries (2012-2018)

|  | Country-Specific EFAs (N=36) | | | | | | | | | Country-Specific CFAs (N=36) | | | | | | | | | | |
| --- | --- | --- | --- | --- | --- | --- | --- | --- | --- | --- | --- | --- | --- | --- | --- | --- | --- | --- | --- | --- |
| **Country** | Push you, shake you, or throw something at you? | Slap you? | Punch with his fist or with something that could hurt you? | Kick you, drag you, or beat you up? | | Try to choke you or burn you on purpose? | | Threaten to attack you with a knife, gun or other weapon? | Twist your arm or pull your hair? | Push you, shake you, or throw something at you? | | Slap you? | Punch with his fist or with something that could hurt you? | | Kick you, drag you, or beat you up? | Try to choke you or burn you on purpose? | | | Threaten to attack you with a knife, gun or other weapon? | Twist your arm or pull your hair? |
| **Central Asia** |  |  |  |  | |  | |  |  |  | |  |  | |  |  | | |  |  |
| Kyrgyz Republic | 0.95 | 0.93 | 0.92 | 0.91 | | 0.95 | | 0.84 | 0.92 | 0.95 | | 0.91 | 0.90 | | 0.93 | 0.94 | | | 0.96 | 0.92 |
| Tajikistan | 0.80 | 0.92 | 0.95 | 0.89 | | 0.92 | | 0.65 | 0.92 | 0.80 | | 0.86 | 0.93 | | 0.92 | 1.02 | | | 0.74 | 0.96 |
| **Latin America and the Caribbean** |  |  |  |  | |  | |  |  |  | |  |  | |  |  | | |  |  |
| Haiti | 0.92 | 0.93 | 0.97 | 0.95 | | 0.70 | | 0.71 | 0.89 | 0.93 | | 0.91 | 0.94 | | 0.92 | 0.88 | | | 0.67 | 0.87 |
| **North Africa, West Asia, Europe** |  |  |  |  | |  | |  |  |  | |  |  | |  |  | | |  |  |
| Armenia | 0.96 | 0.98 | 0.96 | 0.97 | | 0.98 | | 0.98 | 0.95 | 0.99 | | 0.95 | 0.98 | | 0.99 | 0.97 | | | 1.00 | 0.95 |
| Egypt | 0.92 | 0.97 | 0.90 | 0.92 | | 0.86 | | 0.83 | 0.95 | 0.95 | | 0.96 | 0.92 | | 0.92 | 0.90 | | | 0.87 | 0.92 |
| **South and Southeast Asia** |  |  |  |  | |  | |  |  |  | |  |  | |  |  | | |  |  |
| Afghanistan | 0.95 | 0.95 | 0.96 | 0.95 | | 0.89 | | 0.90 | 0.93 | 0.95 | | 0.97 | 0.96 | | 0.93 | 0.89 | | | 0.86 | 0.92 |
| Cambodia | 0.94 | 0.95 | 0.92 | 0.91 | | 0.60 | | 0.80 | 0.88 | 0.96 | | 0.96 | 0.94 | | 0.96 | 0.87 | | | 0.82 | 0.92 |
| India | 0.89 | 0.91 | 0.91 | 0.94 | | 0.86 | | 0.78 | 0.93 | 0.89 | | 0.90 | 0.93 | | 0.92 | 0.83 | | | 0.81 | 0.92 |
| Maldives | 0.95 | 0.96 | 0.98 | | 0.98 | | 0.97 | 0.94 | 0.96 | | 0.96 | 0.94 | | 0.99 | 0.95 | | 0.83 | 0.73 | | 0.91 |
| Myanmar | 0.87 | 0.96 | 0.95 | 0.97 | | 0.81 | | 0.78 | 0.92 | 0.91 | | 0.93 | 0.97 | | 0.91 | 0.78 | | | 0.85 | 0.93 |
| Nepal | 0.89 | 0.93 | 0.94 | 0.96 | | 0.89 | | 0.84 | 0.97 | 0.92 | | 0.97 | 0.97 | | 0.98 | 0.96 | | | 0.84 | 0.96 |
| Pakistan | 0.97 | 0.96 | 0.89 | 0.92 | | 0.86 | | 0.87 | 0.95 | 0.95 | | 0.94 | 0.85 | | 0.98 | 0.98 | | | 0.87 | 0.95 |
| Philippines | 0.91 | 0.91 | 0.94 | 0.97 | | 0.91 | | 0.89 | 0.91 | 0.91 | | 0.86 | 0.89 | | 0.95 | 0.93 | | | 0.82 | 0.92 |
| Timor-Leste | 0.70 | 0.83 | 0.86 | 0.91 | | 0.68 | | 0.66 | 0.92 | 0.76 | | 0.93 | 0.89 | | 0.87 | 0.66 | | | 0.69 | 0.87 |
| **Sub-Saharan Africa** |  |  |  |  | |  | |  |  |  | |  |  | |  |  | | |  |  |
| Angola | 0.87 | 0.92 | 0.92 | 0.93 | | 0.82 | | 0.81 | 0.87 | 0.84 | | 0.91 | 0.94 | | 0.92 | 0.82 | | | 0.78 | 0.88 |
| Benin | 0.91 | 0.87 | 0.94 | 0.97 | | 0.90 | | 0.81 | 0.94 | 0.89 | | 0.88 | 0.93 | | 0.93 | 0.92 | | | 0.93 | 0.91 |
| Burundi | 0.89 | 0.87 | 0.92 | 0.93 | | 0.87 | | 0.84 | 0.90 | 0.89 | | 0.89 | 0.92 | | 0.93 | 0.88 | | | 0.76 | 0.89 |
| Chad | 0.92 | 0.95 | 0.92 | 0.92 | | 0.92 | | 0.85 | 0.94 | 0.89 | | 0.94 | 0.94 | | 0.94 | 0.92 | | | 0.82 | 0.92 |
| Comoros | 0.90 | 0.94 | 0.92 | 0.99 | | 0.88 | | 0.93 | 0.74 | 0.90 | | 0.93 | 0.97 | | 1.01 | 0.82 | | | 0.92 | 0.94 |
| DRC | 0.80 | 0.88 | 0.87 | 0.83 | | 0.87 | | 0.85 | 0.81 | 0.84 | | 0.85 | 0.85 | | 0.85 | 0.91 | | | 0.82 | 0.75 |
| Ethiopia | 0.91 | 0.91 | 0.90 | 0.92 | | 0.89 | | 0.76 | 0.90 | 0.86 | | 0.95 | 0.95 | | 0.93 | 0.80 | | | 0.90 | 0.89 |
| Gabon | 0.84 | 0.80 | 0.95 | 0.94 | | 0.84 | | 0.64 | 0.93 | 0.85 | | 0.88 | 0.98 | | 0.94 | 0.90 | | | 0.86 | 0.91 |
| Gambia | 0.78 | 0.93 | 0.87 | 0.81 | | 0.57 | | 0.96 | 0.86 | 0.83 | | 0.90 | 0.84 | | 0.85 | 0.85 | | | 1.07 | 0.94 |
| Kenya | 0.87 | 0.92 | 0.94 | 0.91 | | 0.85 | | 0.79 | 0.85 | 0.90 | | 0.89 | 0.93 | | 0.93 | 0.85 | | | 0.82 | 0.88 |
| Malawi | 0.86 | 0.94 | 0.90 | 0.90 | | 0.83 | | 0.84 | 0.89 | 0.89 | | 0.90 | 0.92 | | 0.94 | 0.84 | | | 0.83 | 0.89 |
| Mali | 0.79 | 0.83 | 0.83 | 0.88 | | 0.79 | | 0.58 | 0.91 | 0.77 | | 0.84 | 0.85 | | 0.86 | 0.81 | | | 0.58 | 0.74 |
| Mozambique | 0.92 | 0.91 | 0.94 | 0.94 | | 0.89 | | 0.83 | 0.95 | 0.88 | | 0.96 | 0.89 | | 0.91 | 0.84 | | | 0.74 | 0.93 |
| Namibia | 0.93 | 0.94 | 0.96 | 0.98 | | 0.87 | | 0.92 | 0.91 | 0.90 | | 0.97 | 0.98 | | 0.93 | 0.90 | | | 0.86 | 0.89 |
| Nigeria | 0.92 | 0.96 | 0.92 | 0.93 | | 0.88 | | 0.78 | 0.89 | 0.93 | | 0.96 | 0.93 | | 0.93 | 0.83 | | | 0.74 | 0.86 |
| Rwanda | 0.95 | 0.93 | 0.94 | 0.94 | | 0.92 | | 0.83 | 0.89 | 0.91 | | 0.93 | 0.95 | | 0.95 | 0.89 | | | 0.84 | 0.86 |
| Sierra Leone | 0.83 | 0.95 | 0.85 | 0.74 | | 0.82 | | 0.83 | 0.89 | 0.87 | | 0.95 | 0.83 | | 0.74 | 0.76 | | | 0.76 | 0.88 |
| Tanzania | 0.87 | 0.93 | 0.89 | 0.92 | | 0.76 | | 0.76 | 0.81 | 0.88 | | 0.93 | 0.93 | | 0.92 | 0.80 | | | 0.80 | 0.83 |
| Togo | 0.91 | 0.95 | 0.92 | 0.91 | | 0.89 | | 0.82 | 0.89 | 0.92 | | 0.95 | 0.91 | | 0.90 | 0.86 | | | 0.89 | 0.85 |
| Uganda | 0.83 | 0.93 | 0.90 | 0.90 | | 0.85 | | 0.74 | 0.84 | 0.85 | | 0.94 | 0.90 | | 0.91 | 0.85 | | | 0.75 | 0.87 |
| Zambia | 0.86 | 0.90 | 0.89 | 0.91 | | 0.87 | | 0.88 | 0.88 | 0.86 | | 0.87 | 0.86 | | 0.93 | 0.85 | | | 0.79 | 0.90 |
| Zimbabwe | 0.88 | 0.81 | 0.92 | 0.93 | | 0.83 | | 0.77 | 0.88 | 0.86 | | 0.83 | 0.90 | | 0.94 | 0.81 | | | 0.79 | 0.85 |
| Min | 0.70 | 0.80 | 0.83 | 0.74 | | 0.57 | | 0.58 | 0.74 | 0.76 | | 0.83 | 0.83 | | 0.74 | 0.66 | | | 0.58 | 0.74 |
| Max | 0.97 | 0.98 | 0.98 | 0.99 | | 0.98 | | 0.98 | 0.97 | 0.99 | | 0.97 | 0.99 | | 1.01 | 1.02 | | | 1.07 | 0.96 |

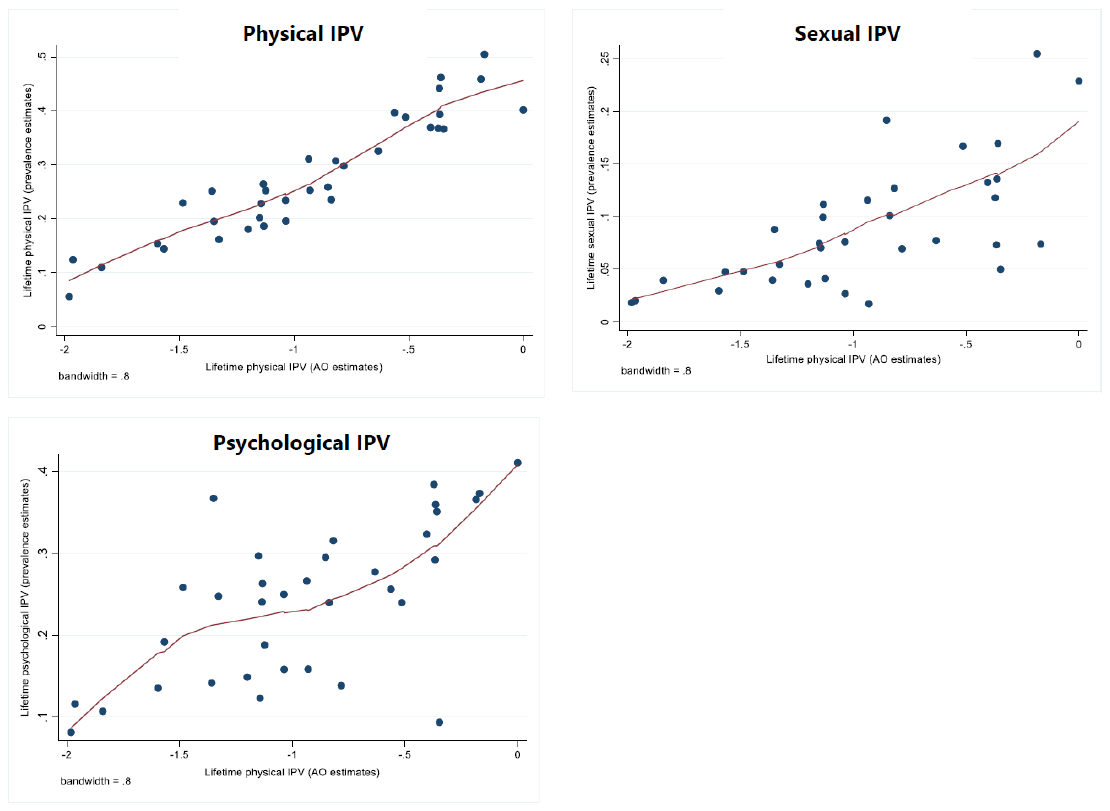

**Figure S1.** Correlations of Prevalence Estimates and Alignment Optimization Estimates of Lifetime Intimate Partner Violence across 35 Countries (outlier removed), 2012-2018.
